# Clinical progression of Parkinson’s disease in the early 21st century: Insights from AMP-PD dataset

**DOI:** 10.1101/2024.01.29.24301950

**Authors:** Mechelle M. Lewis, Xinyi Vivian Cheng, Guangwei Du, Lijun Zhang, Changcheng Li, Sol De Jesus, Samer D. Tabbal, Richard Mailman, Runze Li, Xuemei Huang

## Abstract

**Background:** Parkinson’s disease (PD) therapeutic strategies have evolved since the introduction of levodopa in the 1960s, but there is limited data on their impact on disease progression markers.

**Objective:** Delineate the current landscape of PD progression at tertiary subspecialty care and research centers.

**Method:** Using Accelerating Medicine Partnership-PD (AMP-PD) data harmonized from seven biomarker discovery studies (2010-2020), we extracted: overall [Schwab and England (S&E), PD Questionnaire (PDQ-39)]; motor [Movement Disorders Society Unified PD Rating Scale (MDS-UPDRS)-II and -III and Hoehn & Yahr (HY)]; and non-motor [MDS-UPDRS-I, University of Pennsylvania Smell Identification Test (UPSIT), Montreal Cognitive Assessment (MoCA), and Epworth Sleepiness Scale (ESS)] scores. Age at diagnosis was set as 0 years, and data were tracked for 15 subsequent years.

**Results:** Subjects’ (3,001 PD cases: 2,838 white, 1,843 males) mean age at diagnosis was 60.2±10.3 years and disease duration was 9.9±6.0 years at the baseline evaluation. Participants largely reported independence (S&E, *5y*: 86.6±12.3; *10y*: 78.9±19.3; *15y*: 78.5±17.0) and good quality of life (PDQ-39, *5y*: 15.5±12.3; *10y*: 22.1±15.8; *15y*: 24.3±14.4). Motor scores displayed a linear progression, whereas non-motor scores plateaued ∼10-15 years. Younger onset age correlated with slower overall (S&E), motor (MDS-UPDRS-III), and non-motor (UPSIT/MoCA) progression, and females had better overall motor (MDS-UPDRS-II-III) and non-motor (UPSIT) scores than males.

**Conclusions:** Twenty-first century PD patients remain largely independent in the first decade of disease. Female and young age of diagnosis were associated with better clinical outcomes. There are data gaps for non-whites and metrics that gauge non-motor progression for >10 years after diagnosis.

## Introduction

In 1817, James Parkinson described the clinical syndrome “Shaking Palsy”^1^ that bears his name. In the following century, Parkinson’s disease (**PD**) was linked to progressive neuronal loss in the substantia nigra (**SN**)^2,3^ and presence of Lewy bodies. The key driving pathophysiological change was thought to be the continual decrease in dopaminergic function.^4,5^ This is evident by pathological changes including α-synuclein (**αSyn**)-positive Lewy bodies/neurites and dopamine neuron loss in the substantia nigra pars compacta (**SNc**) of the basal ganglia (**BG**).^6,7^ The clinical presentation of motor dysfunction (tremor, rigidity, bradykinesia) represents the threshold when the system can no longer accommodate continued loss of dopaminergic innervation/tone. Without effective treatment, PD patients previously showed rapid disease progression, became significantly disabled by ∼5 years, and died on average ∼9.4 years after diagnosis.^8^ By the early 1960s, ascertainment of the dopaminergic nigrostriatal pathway of the BG transformed our understanding of PD pathophysiology.^9^ The availability of levodopa effectively restored lost dopaminergic tone^5^ and led to a new era for PD patients, with substantially improved symptoms and lengthened lifespan.^8,10-15^

Over the past half century, many newer therapies were added to enhance/normalize nigrostriatal dopaminergic functions. They include enzyme inhibitors [i.e., against monoamine oxidase (MAO) or catecholamine O-methyl transferase (COMT)] that reduce dopamine and levodopa metabolism, dopamine agonists that directly activate post-synaptic dopamine D2/3 receptors (e.g., pramipexole, ropinirole, and rotigotine), and neurosurgical procedures [pallidotomy and deep brain stimulation (DBS)]. Despite these advances, levodopa remains the cornerstone for PD treatment, and responsiveness to levodopa is an integral part of clinical diagnosis and selecting PD candidates for DBS surgery. Yet even with optimal therapy, PD progresses with worsening motor and non-motor symptoms that contribute to the deterioration of patient social functioning and general quality of life.

For this reason, the past four decades of PD research focused on disease-modifying interventions to slow, arrest or “cure” PD progression.^16-23^ A failure to discover an effective neuroprotective drug^24^ helped propel research efforts to identify disease biomarkers that would both suggest new leads and also allow for quantitative assessment of clinical efficacy.^16^ Examples of such biomarkers include radioligand-based imaging techniques reflecting nigrostriatal integrity (PET and SPECT),^25^ MRI methods to assess the state of the SN,^26-31^ and αSyn fluid-based assays^32-35^ to measure different aspects of PD pathology. The current study was designed to help understand the needs of the PD patient community by evaluating PD clinical progression under the current state of best practices at tertiary subspecialty care and research centers.

## Methods

### Source data: Accelerating Medicine Partnership in PD (AMP-PD)

The Accelerating Medicine Partnership PD (AMP-PD; https://amp-pd.org/about^36^) was created to harmonize biomarker-related datasets from different PD cohorts and make them available to the research community. This study used *Release 2.0* that included: the Parkinson’s Progression Marker Initiative (PPMI); the Fox Investigators for New Discovery of Biomarkers in PD (BioFIND); the PD Biomarkers Program [PDBP, sponsored by the National Institute of Neurological Disorders and Stroke (NINDS)]; the Harvard Biomarker Study (HBS); the International Lewy Body Dementia Genetics Consortium Genome Sequencing in Lewy body dementia case-control cohort (LBD); the MJFF LRRK2 Cohort Consortium (LCC); and the Study of Isradipine as a Disease Modifying Agent in Subjects With Early Parkinson Disease, Phase 3 (STEADY-PD3) cohorts. All data in the AMP-PD dataset are deidentified. All participants fully consented to broad clinical data sharing and were informed of how data may be used.

### Extracting PD status and demographic information at baseline

We retrieved data of participants in the AMP-PD database (Release v2.0) as of December 16, 2021. Controls (n=4,355) and participants who carried a diagnosis other than idiopathic PD (n=2,828) were excluded. Our final dataset (N=3,001) included all subjects diagnosed as “idiopathic PD” or “Parkinson’s disease” at the screening or baseline visit (Figure 1). Four patients had repeated records containing contradictory demographic information, so we selected their most recent data. Age of participants, gender, education, and ethnicity/race were provided by the Demographics file at baseline. The age at PD diagnosis was extracted from the baseline PD Medical History form. Disease duration at baseline was defined as the difference between age at PD diagnosis and age at baseline (enrolled in the studies).

**Figure 1:**
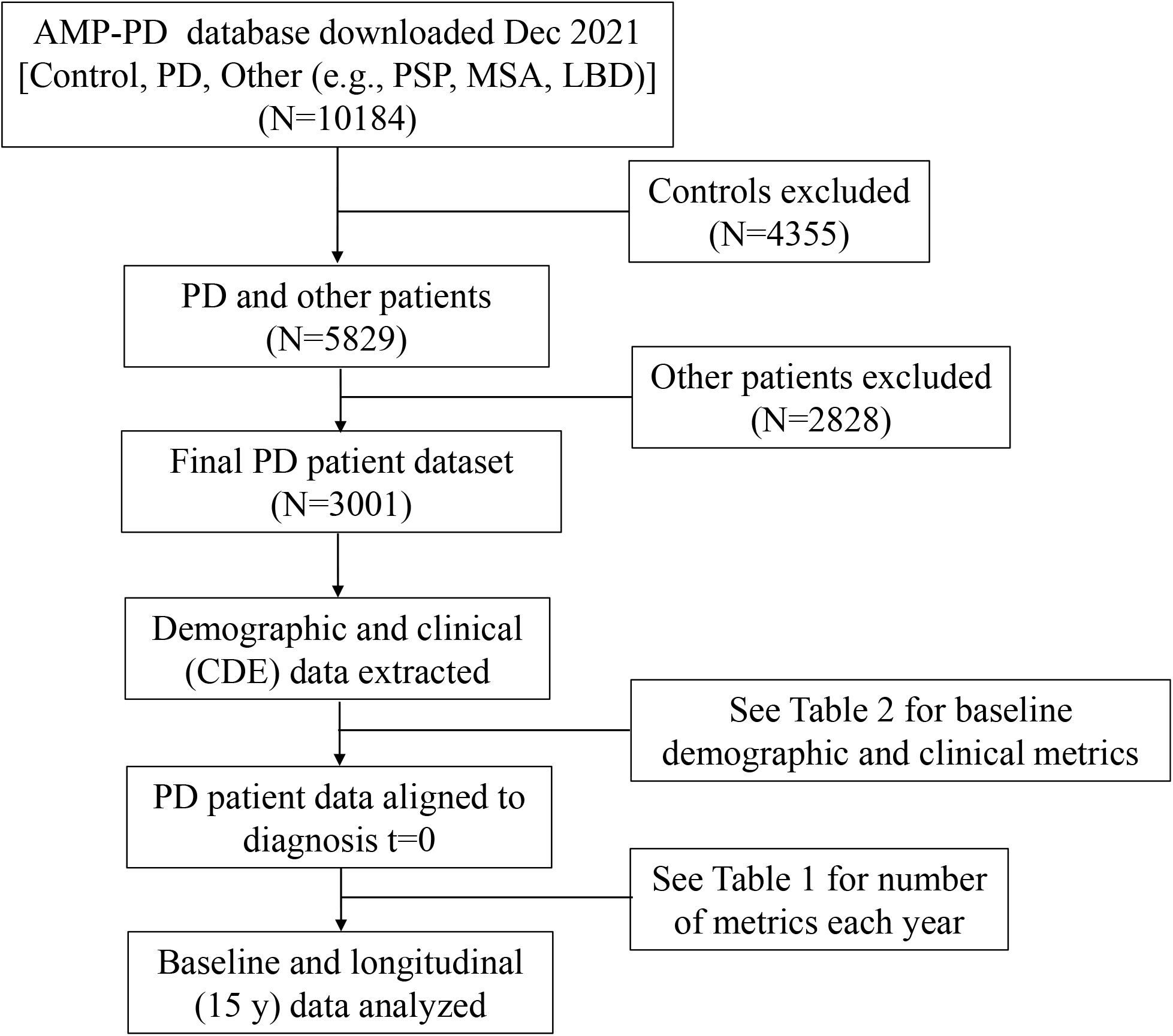
Summary of the study design. Data from the AMP-PD database were downloaded and included information from both control and PD participants. Controls (n=4,355) and participants who carried a diagnosis other than idiopathic PD (n=2,828) were excluded.

### Extracting clinical metrics of interest for disease status and progression

Clinical data were designed to be collected at baseline and every subsequent 6-month epoch for each biomarker study. All PD patients in the AMP-PD dataset have at least one of the following NINDS-defined common data elements (CDE) for PD.

### Schwab & England (S&E) Activities of Daily Living (ADL) scale

This scale^37^ measures percentage of independence on a scale of 0-100%, with 100% representing complete independence and 0% fully-dependent.

### PD Questionnaire (PDQ-39)

The PD questionnaire (PDQ)-39 is a 39-item self-report questionnaire^38^ assessing PD-specific health-related quality of life in eight domains. Five options (never, occasionally, sometimes, often, or always) evaluate each item over the past month. The maximum score is 100.

### Hoehn and Yahr (H&Y) scale

H&Y score^8^ tries to describe progression of PD symptoms. It ranges from I-V with I=unilateral symptoms, II=bilateral symptoms without balance impairment, III=bilateral symptoms with impaired postural reflexes, IV=severe disabling disease but still able to stand/walk unassisted, V=bed or wheelchair bound.

### Movement Disorder Society Unified PD Rating Scale (MDS-UPDRS)

The MDS-UPDRS^39^ consists of four subscales evaluating non-motor aspects of daily living (MDS-UPDRS-I, maximum total score=52), motor aspects of daily living as reported by subjects and/or caregiver (MDS-UPDRS-II, maximal total score=52), motor symptoms assessed by a trained examiner (MDS-UPDRS-III, maximal total=132), and motor complications (MDS-UPDRS-IV, maximum total score=24). Our current study evaluated subscores I, II, and III.

### University of Pennsylvania Smell Identification Test (UPSIT)

The UPSIT^40^ assesses olfactory function using 40 scratch and sniff items (total score=40). Items list four multiple choice options for participants to select the correct odor associated with each scratch panel.

### Montreal Cognitive Assessment (MoCA)

MoCA^41^ is a short assessment of overall cognition. With a maximum score=30, it evaluates different cognitive domains including attention and concentration, executive function, memory, language, visuo-constructive skills, conceptual thinking, calculations, and orientation.

### Epworth Sleepiness Scale (ESS)

The ESS^42^ is a subjective measurement of a person’s daytime sleepiness. It describes eight scenarios and asks participants to rate their tendency to become sleepy: 0 (no chance of dozing off) to 3 (high chance of dozing off). Maximum total score is 24, with scores >10 reflecting increased sleepiness.

Higher H&Y, MDS-UPDRS, PDQ-39, and ESS scores reflect worse function, whereas higher Schwab and England, UPSIT, and MoCA scores reflect better function.

#### Statistical analysis

Each AMP-PD cohort includes patients with different disease durations at baseline. Thus, we set the time of diagnosis as time = 0 (t0) years. Although the AMP-PD dataset harmonized data from 2010-2020, our strategy allowed us to ascertain PD progression over 15 years since some participants had disease duration >10 years. We then identified the number of CDE metrics at each timepoint, as well as disease duration, gender, education, ethnicity, and race. We divided participants into three groups based on their age at diagnosis: <50; 50-70; and >70 y. We computed the average and standard deviation (SD) for each CDE at 0, 5, 10, and 15 years of disease. We evaluated CDE progression using partial linear varying coefficient models that included effects of age at diagnosis and gender. Since patients had different numbers of follow-up visits and different durations of observation, we designed and implemented a novel statistical learning strategy to leverage all AMP-PD data and handle missing data using the following three steps:

##### Step 1

We determined the variables and how many years of data to include in the model. We assessed whether each variable was balanced between subgroups. For each disease duration, we examined the adequacy of data points and determined a cutoff value. For the variables included in the model, we determined whether to study their dynamic or fixed effect. We explored simple linear regression results and boxplots of the score distribution at each disease duration (data not shown). If we observed significant differences in a metric across years, it was treated as a dynamic variable.

##### Step 2

We explored the dynamic nature and effects of age at diagnosis and gender over 15 y of disease progression using partial linear varying coefficient models. Although multiple linear regression could be adopted to study variable impacts and their interactions, we elected to limit our study to linear interactions knowing that this may not capture potential non-linear trends. To overcome this limitation, we chose to relax the linear assumptions of regression to explore the non-linear effects of covariates that may vary with disease progression. In our current study, age was captured as a fixed linear effect and gender as a varying effect. Therefore, we applied a partial linear varying coefficient model (PLVCM) for each metric with age and gender as covariates.

We adopted a B-spline regression for PLVCM to capture the dynamic nature of disease progression. We considered the relationship among the scores *(Y)*, and gender (*X*_*1*_)and age (*X*_2_). We included an intercept term also varying with disease duration to capture the nonlinear trend. Gender also varied with disease duration. We centered age to its means to control the effect of scaling. With *t* denoting disease duration, the PLVCM model was as follows:

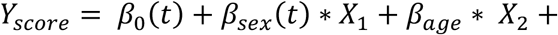

This model was used to fit each clinical score. Each metric with confidence intervals was graphed over the 15-year span.

##### Step 3

Based on previous literature^29,31,43-45^ and our PLVCM results, we conducted linear mixed effect regression (LMER) analyses on three PD subgroups defined based on disease duration: 0-5 y, 6-10 y, and 11-15 y. We considered a random effect for each patient (*ϵid*), and a fixed effect for disease duration (*X*_*1*_)and the covariates of age (*X*_2_)and gender (*X*_3_).We also included the interaction between disease duration and gender (*X*_*1*_\**X*_3_)if it was indicated by the results in the preceding analyses. An interaction term was included if the PLVCM lines for the two genders were discernibly unparallel. The LMER model assumed the following form:

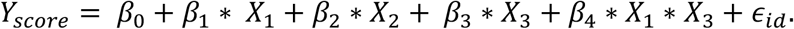

## Results

### Sample size and baseline demographic and clinical features of AMP-PD dataset

For these subjects (Table 1), limited data points (N≤119) were observed for PD patients with disease duration >10 y. Of the 3,001 patients, the mean age at diagnosis [± SD] was 60.2±10.3 y, with ca. 60% of patients (n=1,843) being male and 1,158 female (Table 2). Most had an age at diagnosis between 50-70 y (n=1972), whereas 211 were diagnosed <50 years old and 448 >70 years old. The mean disease duration was 9.9±6.0 y at the baseline evaluation. The majority had disease duration 0-5 y (n=1,915) at baseline, with fewer subjects having longer disease duration (disease duration 5-10 y N=541, 11-15 y n=254, >15 y n=163). Most subjects had an education level of 12-16 y (n=1,842), followed by advanced education of >16 y (n=811), <12 y (n=201), unknown (n=146), and 0 y (n=1). Ethnicity and race were skewed heavily toward non-Hispanic (n=2,603) and White (n=2,838) subjects.

**Table 1.** Total number of AMP-PD PD participants per year for each clinical metric.

| Year | 0 | 1 | 2 | 3 | 4 | 5 | 6 | 7 | 8 | 9 | 10 | 11 | 12 | 13 | 14 | 15 | >15 |
| --- | --- | --- | --- | --- | --- | --- | --- | --- | --- | --- | --- | --- | --- | --- | --- | --- | --- |
| **Functional scales** |  |  |  |  |  |  |  |  |  |  |  |  |  |  |  |  |  |
| Schwab & England stage | 495 | 707 | 778 | 760 | 715 | 678 | 632 | 536 | 379 | 239 | 171 | 121 | 82 | 79 | 75 | 66 | 54 |
| PDQ-39 | 119 | 168 | 194 | 212 | 178 | 167 | 148 | 125 | 107 | 95 | 67 | 60 | 42 | 43 | 41 | 41 | 28 |
| **Motor scales** |  |  |  |  |  |  |  |  |  |  |  |  |  |  |  |  |  |
| Hoehn & Yahr stage | 644 | 802 | 853 | 825 | 748 | 65 | 616 | 529 | 367 | 232 | 160 | 113 | 78 | 61 | 66 | 63 | 41 |
| MDS-UPDRS-II | 491 | 699 | 762 | 746 | 690 | 653 | 591 | 505 | 345 | 215 | 132 | 101 | 68 | 56 | 58 | 58 | 39 |
| MDS-UPDRS-III | 497 | 713 | 773 | 756 | 700 | 664 | 595 | 501 | 346 | 215 | 135 | 101 | 68 | 56 | 58 | 58 | 39 |
| **Non-Motor scales** |  |  |  |  |  |  |  |  |  |  |  |  |  |  |  |  |  |
| MDS-UPDRS-I | 491 | 699 | 763 | 746 | 691 | 653 | 591 | 505 | 345 | 215 | 132 | 101 | 68 | 56 | 58 | 58 | 39 |
| UPSIT | 450 | 374 | 297 | 272 | 232 | 225 | 239 | 268 | 240 | 166 | 116 | 92 | 70 | 73 | 66 | 61 | 50 |
| MoCA | 502 | 722 | 784 | 771 | 721 | 679 | 628 | 530 | 377 | 238 | 165 | 120 | 82 | 76 | 74 | 66 | 54 |
| ESS | 454 | 673 | 751 | 732 | 681 | 633 | 597 | 507 | 356 | 219 | 136 | 95 | 57 | 57 | 55 | 49 | 42 |

Abbreviations: ESS = Epworth Sleepiness Scale; MoCA = Montreal Cognitive Assessment; PDQ-39 = 39-item Parkinson's Disease Questionnaire; UPDRS-I/II/III = Unified Parkinson's Disease Rating Scale – subscales of non-motor activities reported by patients (I), motor activities reported by patients (II), and motor symptoms assessed by rater exam (III); UPSIT = University of Pennsylvania Smell Identification Test.

**Table 2.** Baseline demographic and clinical characteristics of AMP-PD participants.

| **Demographic characteristics (N=3001 total)** |  |  |  |
| --- | --- | --- | --- |
| **Age at diagnosis (years)** | **N** | **Gender** | **N** |
| Mean | 60.2±10.3 | Male | 1843 |
| < 50 | 436 | Female | 1158 |
| 50-70 | 1972 | Missing | 0 |
| >70 | 448 |  |  |
| Missing | 145 |  |  |
| **Disease duration at baseline (in years)** |  | **Ethnicity** |  |
| Mean | 9.9±6.0 | Hispanic-Latino | 129 |
| 0-5 | 1915 | Not Hispanic-Latino | 2603 |
| 6-10 | 541 | Unknown | 266 |
| 11-15 | 254 | Missing | 3 |
| >15 | 163 |  |  |
| Missing | 118 |  |  |
| **Education (in years)** |  | **Race** |  |
| 0 | 1 | White | 2838 |
| <12 | 201 | Black or African American | 39 |
| 12-16 | 1842 | Asian | 32 |
| >16 | 811 | Other* | 43 |
| Unknown | 146 | Missing | 49 |
| **Clinical characteristics (mean ± SD)** |  |  |  |
| **Functional** |  | **Non-motor** |  |
| Schwab and England stage | 87.1±12.8 | MDS-UPDRS-I | 8.7±5.9 |
| PDQ-39 | 16.6±13.1 | UPSIT | 20.3±8.3 |
| **Motor** |  | **MoCA** | 26.0±3.6 |
| Hoehn & Yahr stage | 1.9±0.6 | Epworth Sleepiness Scale | 7.4±4.6 |
| MDS-UPDRS-II | 9.5±7.0 |  |  |
| MDS-UPDRS-III | 24.4±12.6 |  |  |

Error terms are standard deviations.
\*Other includes American Indian/Alaska Native, Native Hawaiian/Other Pacific Islander, Arab, Other, Unknown.
Abbreviations: MoCA = Montreal Cognitive Assessment; PDQ-39 = 39-item Parkinson's Disease Questionnaire; Schwab and England = Schwab and England Activities of Daily Living; UPDRS-I/II/III = Unified Parkinson's Disease Rating Scale – subscales of non-motor activities reported by patients (I), motor activities reported by patients (II), and motor symptoms assessed by rater exam (III); UPSIT = University of Pennsylvania Smell Identification Test.

### Clinical progression over 15 years

#### Overall function

The average S&E score at baseline was 87.1±12.8% (Table 2). S&E scores worsened steadily until ∼12 y and then plateaued at a score of ∼75% (Figure 2A), consistent with the percept of patients in various H&Y stages as it relates to ADL. The mean ± SD scores at 5, 10, and 15 y were 86.6±12.3%, 78.9%±19.3%, and 78.8±17.0%, respectively. The average PDQ-39 score was 16.6±13.1 at baseline. PDQ-39 scores worsened across the disease course (Figure 2B), although variance was relatively high perhaps due to the small sample size in later years. The mean ± SD PDQ-39 scores were 15.5±12.3 at 5 y, 22.1±15.8 at 10 y, and 24.3±14.4 at 15 y.

**Figure 2:**
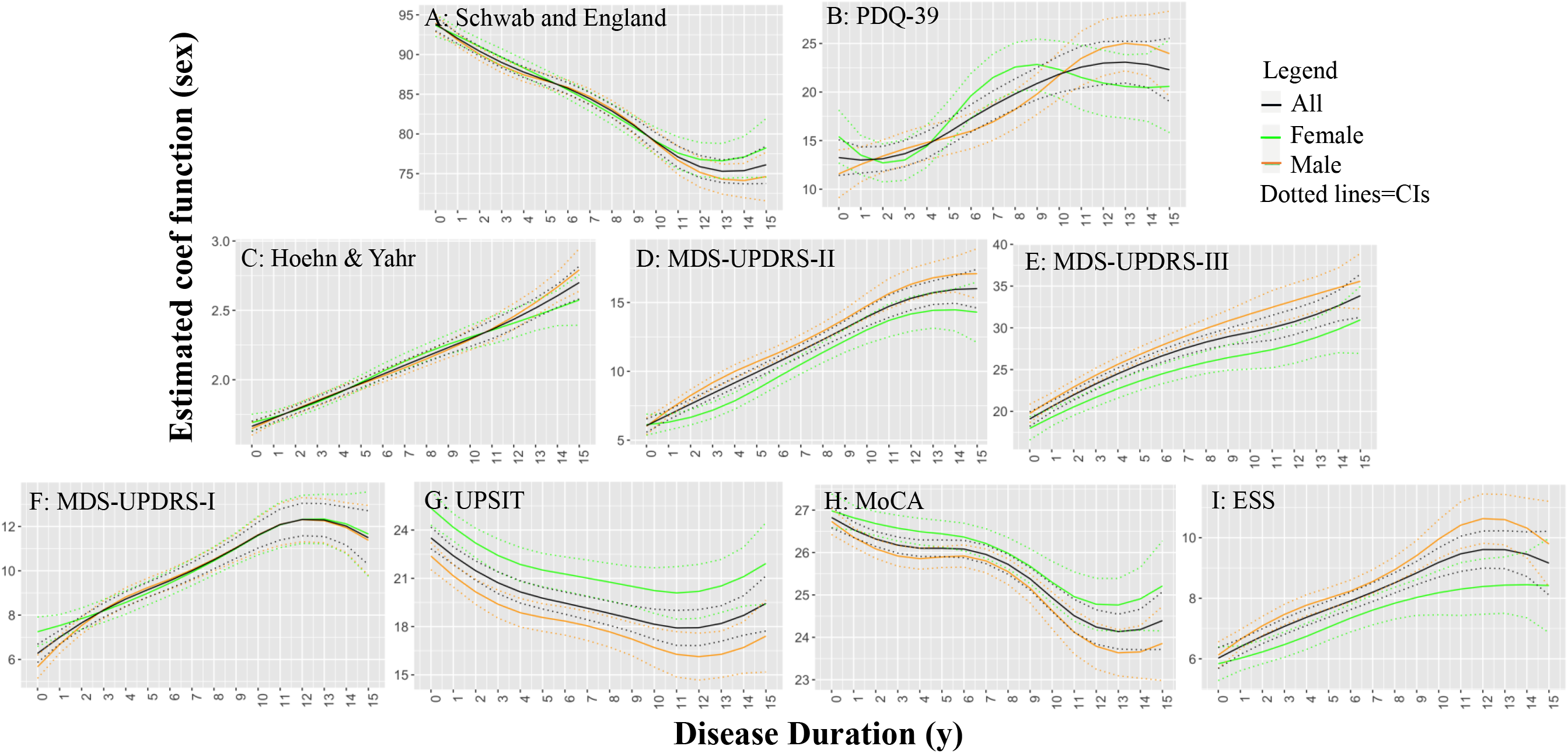
Progression of PD clinical metrics over 15 years displayed by sex distribution. Progression of clinical metrics for the PD cohort over 15 years (all, black solid lines), with estimated confidence intervals (dotted lines). In addition, gender effects for each year (solid lines, females represented in green and males in orange) are shown with estimated confidence intervals (dotted lines). <u>Functional scales:</u> (A) Schwab & England scale, (B) 39-item PD Questionnaire (PDQ-39); <u>Motor scales:</u> (C) Hoehn & Yahr, Movement Disorders Society Unified PD Rating Scale (MDS-UPDRS) subscales representing (D) motor activities reported by patients (II), and (E) motor symptoms assessed by rater exam (III); <u>Non-motor scales:</u> (F) MDS-UPDRS-I representing non-motor activities reported by patients, (G) University of Pennsylvania Smell Identification Test (UPSIT), (H) Montreal Cognitive Assessment (MoCA), and (I) Epworth Sleepiness Scale (ESS) scores.

#### Motor function

H&Y stage and MDS-UPDRS motor (-II and -III) scores captured disease progression linearly across 0-15 y (Figure 2C-F). H&Y scores were 2.0±0.6 at 5 y (bilateral disease without postural instability), 2.3±0.7 at 10 y, and 2.6±0.8 at 15 y (bilateral disease with some postural instability), reflecting a gradual progression to postural instability. MDS-UPDRS motor subscores were consistent, showing that PD patients were doing relatively well over 15 y despite gradual worsening of their motor symptoms and scores. Specifically, MDS-UPDRS-II scores were 10.0±6.8 at 5 y, 13.9±9.0 at 10 y, and 16.0±8.7 at 15 y, and MDS-UPDRS-III scores were 26.1±12.2 at 5 y, 28.7±15.8 at 10 y, and 31.1±17.1 at 15 y.

#### Non-motor function

MDS-UPDRS non-motor (I) scores captured disease progression in the first 10 y, but plateaued at ∼11 y (Figure 2F): mean (±SD) scores were 9.3±5.9 at 5 y, 12.5±7.2 at 10 y, and 11.6±5.9 at 15 y. PD patients has an average UPSIT score of 20.3±8.3, indicative of severe anosmia at baseline.^46^ UPSIT scores showed an almost linear worsening over 0-8 y and then plateaued (Figure 2G) with wide variance [20.3±8.4 at 5 y, 19.5±7.4 at 10 y, and 20.7±8.2 at 15 y (mean (±SD)]. MoCA scores were 26.0±3.6 at baseline. They averaged 26.2±3.5 at 5 y, 25.0±4.2 at 10 y, and 24.9±4.5 at 15 y, indicating a gradual progression to mild cognitive impairment by year 10 (Figure 2H). ESS scores were 7.4±4.6 at baseline, and increased linearly in years 0-10 when they appeared to level off at an average score of ∼9 (Figure 2I), with all in normal range.^42^

### Effect of gender on clinical progression

The effect of gender and age in each disease epoch (0-5 y, 6-10 y, and 11-15 y) are shown in Supplemental Table 1.

Overall function: S&E scores were similar for males and females across the duration of disease observation (Figure 2A). For PDQ-39, scores for the year 6-9 epoch for females were slightly (but not significantly) worse than those of males (Figure 2B).

Motor function: Females and males progressed similarly on the H&Y scale (Figure 2C). Females initially had similar MDS-UPDRS-II and -III scores, however, they had significantly better scores than males after year 1 (all p < 0.015) (Figures 2D and 2E).

Non-motor function: Female participants initially had worse MDS-UPDRS-I scores but only for years 0 and 1 (all p<0.002), with no difference thereafter (Figure 2D). Females had significantly better UPSIT scores than males across the 0-15 years (all p<0.035), with a ∼2.5-point difference initially, and a ∼4.2-point difference in the later disease course (Figure 2G). Females initially had significantly better MoCA scores than males (years 0-1, p<0.048; Figure 2H). They also tended to have better MoCA scores than males throughout the disease course but were no longer significant after year 3 (all p>0.05). Females and males initially had similar ESS scores, but then male scores rose significantly at year 2 and were higher than females throughout the remaining disease course (all p<0.038, Figure 2I).

### Effect of age at diagnosis on clinical progression

Age at diagnosis affected all clinical metrics (Figure 3 and Supplemental Table 2), with younger age associated with slower progression.

**Figure 3:**
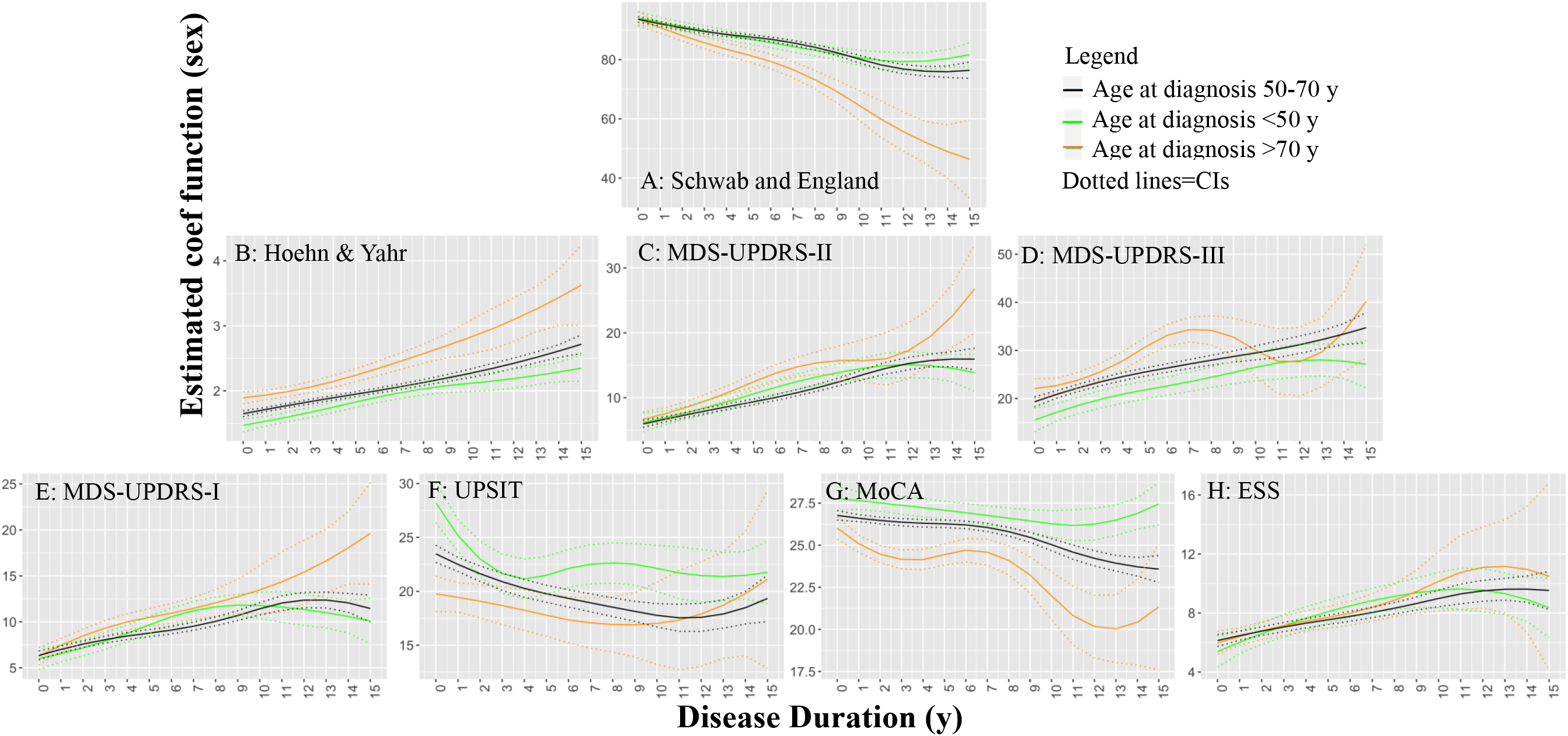
Effect of diagnosis age on PD progression. Progression of clinical metrics for the PD cohort over 15 years (all, black solid lines), with estimated confidence intervals (dotted lines). In addition, age effects for each year (solid lines, age at diagnosis <50 y represented in green and >70 y in orange) are shown with estimated confidence intervals (dotted lines). <u>Functional scales:</u> (A) Schwab & England scale; <u>Motor scales:</u> (B) Hoehn & Yahr, Movement Disorders Society Unified PD Rating Scale (MDS-UPDRS) subscales representing (C) motor activities reported by patients (II), and (D) motor symptoms assessed by rater exam (III); <u>Non-motor scales:</u> (E) MDS-UPDRS-I representing non-motor activities reported by patients, (F) University of Pennsylvania Smell Identification Test (UPSIT), (G) Montreal Cognitive Assessment (MoCA), and (H) Epworth Sleepiness Scale (ESS) scores. PDQ-39 scores are not shown in this figure due to the lack of sufficient data points, particularly after year 6.

#### Overall function

S&E scores for PD patients diagnosed at age <50 or 50-70 y worsened steadily over 10 y and then plateaued (Figure 3A), whereas those diagnosed at age >70 y declined slowly until year 7, thereafter decreasing more rapidly (age at diagnosis effect β=-0.151±0.015, p<0.001). Due to the limited number of PDQ-39 values after year 10, the effect of age for PDQ-39 was not evaluated.

#### Motor function

H&Y scores steadily worsened for all PD patients until year 5. After year 5, those diagnosed at age >70 y increased significantly more than those diagnosed at age <50 or 50-70 y (Figure 3B, age at diagnosis effect β=0.014±0.001, p<0.001). MDS-UPDRS-II scores increased in all groups similarly until year 12 when scores for those diagnosed >70 increased significantly more (Figure 3C, age at diagnosis effect β=0.038±0.09, p<0.001). MDS-UPDRS-III scores were significantly lower in PD patients diagnosed at age <50 y compared to those diagnosed at age >70 y (β=0.219±0.016, p<0.001) until year 9 (Figure 3D).

#### Non-motor function

MDS-UPDRS-I scores were similar in all age groups over 0-10 y. After year 10, the scores significantly increased in PD patients diagnosed at age >70 y compared to other groups (Figure 3E, group effect β=0.029±0.007, p<0.001). UPSIT scores in patients diagnosed at age >70 y were relatively stable over 15 y (Figure 3F), whereas those diagnosed at age <50 y showed a rapid decline in scores in years 0-3. Patients diagnosed at age 50-70 y had a gradual decline in UPSIT scores until year 10 when they plateaued (group effect β=-0.163±0.015, p<0.001). MoCA scores declined gradually over 10 y and then plateaued for age <50 and 50-70 y groups. For the age >70 y group, MoCA scores decreased in 0-3 years and decreased steeply again in years 9-12 (Figure 3G, β=-0.104±0.004, p<0.001). ESS scores increased similarly in years 0-9 for all age groups. At year 12, age >70 group increased faster than other age groups (Figure 3H, age at diagnosis effect β=-0.013±0.006, p=0.029).

## Discussion

We evaluated the progression of clinical measures over the first 15 years using the AMP-PD dataset generated by tertiary subspecialty care and research centers between 2010-2020. We found that PD patients remained largely independent in the first decade of their disease. Female gender and young age of onset were associated with a better clinical course. These data fill in some of the knowledge gap in PD progression for early 21^st^-century patients. There are, however, data gaps regarding non-white individuals and for metrics that gauge non-motor progression for those with disease duration >10 years.

Our analysis suggests that PD patients remained largely independent with a good quality of life in the first 15 years of disease. The exception, not surprisingly, was for patients with onset of illness after age 70 y who tended to lose independence after 10 years of illness as reflected by the sharp worsening in their S&E scores (Figure 3A). This may be due partly to their worsening gait impairment as reflected by an increase in their H&Y score to ≥3 (Figure 3B), as well as worsening cognition indicated MoCA scores <23 (Figure 3G) and increased daytime sedation (ESS score >10, Figure 3H). Overall, MoCA scores dropped from 27 to 25 (out of a total of 30) by year 10 indicating a gradual progression to mild cognitive impairment. In subjects with younger onset of disease, MoCA scores were relatively stable over the course of 15 years, but in subjects with disease onset >70 years, MoCA scores continued to worsen through the course of the disease, although they seemingly stabilized between year 3 and year 8 of the disease (Figure 3G).

It is known that female sex is associated with a lower risk of developing PD, although the exact mechanism is unknown despite many factors having been suggested.^47-50^ Females also may experience less rigidity,^51^ with postural instability and levodopa-induced motor complications^52^ emerging later than in males (although with a higher propensity). Consistent with these prior reports, female PD participants in the AMP-PD dataset overall had better motor (MDS-UPDRS-II and -III) and non-motor (cognition, olfaction, sleepiness) scores than males. The differences persisted over years 0-10, or in the case of the UPSIT, were even greater. These new data extend previous studies that evaluated PD progression over a more limited time frame (4-8 y),^53,54^ and are consistent with reports of female PD patients experiencing a milder form of disease.^51,52^

Consistent with one prior report,^55^ we found that both females and males with PD had similar declines in cognitive scores (MoCA) after 7 years. Conversely, others have reported that males have greater cognitive impairment and more rapid progression than females.^56,57^ Our report was based only on MoCA scores over an epoch (15 years), whereas these latter studies^56,57^ utilized a broad battery of neurocognitive tests and shorter follow-up period. Thus, the discordance between our data and previous studies may suggest that males are more affected by changes to specific high-level tasks, but that changes in routine cognitive function is affected similarly in both sexes. This issue is worthy of future investigation.

The impact of age of onset has been studied frequently,^58-60^ and younger age at PD diagnosis has been associated with decreased clinical severity^59,61^ and slower progression.^60,62^ Our analysis is consistent with this literature, except that UPSIT scores in patients diagnosed at age <50 years showed a more rapid decline in years 0-3 that would not be predicted by other measures. This may be since younger PD patients have a better sense of smell at baseline (Figure 3F) and reflect they have more brain reserve at that time. The rapid decline after initial diagnosis is puzzling and worthy of further investigation. It was noted that younger-age-of-onset patients also may have more resilience in dealing with a non-fatal illness, practice lifestyles that modify neurodegenerative processes (e.g., meditation), and/or enrich their environment through education or other activities (see^63^ for review). Conversely, young-onset PD patients may develop levodopa-induced dyskinesias and motor fluctuations earlier than older-onset patients,^64,65^ but whether this reflects a biological difference^60^ versus method of treatment is unclear.

## Conclusions

This study provides new insights into the current state of PD clinical disease progression and novel opportunities for future clinical and basic science research. Our analysis suggests that younger-age-of-onset and female sex are associated with lesser disease severity. Importantly, current standard-of-care approaches for these 21^st^-century PD patients allow them to be largely independent with relatively good quality of life in the first 10 years of disease. It also was clear that there are two significant gaps in the available data. One of these is the paucity of data for non-white patients. It is very important to determine if other populations have the same trajectories and effects. Equally important is the need to develop and apply clinical metrics (particularly non-motor ones) longitudinally for gauging disease beyond 10 years. There is little data on patients with very advanced/late-stage PD (i.e., disease duration >10 years). With recent data suggesting new classes of drugs may be useful in later stages of disease,^66-68^ sensitive metrics that can quantify activities of daily living and quality of life in this population is a priority, especially if it reflects the perspective of both patients and caregivers.

## Data Availability

All data produced in the present work are contained in the manuscript

## Acknowledgement

Data used in the preparation of this article were obtained from the Accelerating Medicine Partnership® (AMP®) Parkinson’s Disease (AMP PD) Knowledge Platform. The AMP® PD program is a public-private partnership managed by the Foundation for the National Institutes of Health and funded by the National Institute of Neurological Disorders and Stroke (NINDS) in partnership with the Aligning Science Across Parkinson’s (ASAP) initiative; Celgene Corporation, a subsidiary of Bristol-Myers Squibb Company; GlaxoSmithKline plc (GSK); The Michael J. Fox Foundation for Parkinson’s Research; Pfizer Inc.; Sanofi US Services Inc.; and Verily Life Sciences. ACCELERATING MEDICINES PARTNERSHIP and AMP are registered service marks of the U.S. Department of Health and Human Services. For up-to-date information on the study, visit https://www.amp-pd.org.

## Author contributions

1. Research project: A. Conception, B. Organization, C. Execution;
2. Statistical Analysis: A. Design, B. Execution, C. Review and Critique;
3. Manuscript: A. Writing of the first draft, B. Review and Critique; Mechelle M. Lewis: 1A, 1B, 2C, 3A, 3B. Xinyi Vivian Cheng: 1B, 1C, 2B, 2C, 3A, 3B. Guangwei Du: 1A, 1B, 1C, 2A, 2B, 3B. Lijun Zhang: 1C, 3B. Changcheng Li: 1C, 3B. Sol De Jesus: 2C, 3B. Samer Tabbal: 2C, 3B. Richard Mailman: 1A, 2C, 3B Runze Li: 1A, 1B, 1C, 2A, 2C, 3B. Xuemei Huang: 1A, 1B, 1C, 2A, 2C, 3B.

## References

1. Parkinson J. An Essay on the Shaking Palsy. London: Sherwood, Neely and Jones, 1817.

2. Ehringer H, Hornykiewicz O. Verteilung vn Noradrenalin und Dopamin (3-hydroxytyramine) in Gehirn des Menshen und ihr Verhalten bei Evkrankungen des extrapyramidalen systems [Distribution of noradrenaline and dopamine (3-hydroxytyramine) in the human brain and their behavior in diseases of the extrapyramidal system]. Klin Wochenschr 1960;38:1236–1239.

3. Hornykiewicz O. Die Topische Lokalisation und das verhalten von noradrenalin und dopamine (3-Hydroxytyramin) in der substantia nigra des normalen und parkinsonkranken menschen. WienKlinWochenschr 1963;75:309–312.

4. Erro R, Picillo M, Vitale C, et al. The non-motor side of the honeymoon period of Parkinson’s disease and its relationship with quality of life: a 4-year longitudinal study. Eur J Neurol 2016;23(11):1673–1673.

5. Rascol O, Payoux P, Ory F, Ferreira JJ, Brefel-Courbon C, Montastruc JL. Limitations of current Parkinson’s disease therapy. Ann Neurol 2003;53 Suppl 3:S3–12; discussion S12-15.

6. Calne DB, Snow BJ, Lee C. Criteria for diagnosing Parkinson’s disease. Ann Neurol 1992;32 Suppl:S125–S127.

7. Hughes AJ, Daniel SE, Kilford L, Lees AJ. Accuracy of clinical diagnosis of idiopathic Parkinson’s disease: a clinico-pathological study of 100 cases. J Neurol Neurosurg Psychiatry 1992;55(3):181–181.

8. Hoehn MM, Yahr MD. Parkinsonism: onset, progression and mortality. Neurology 1967;17(5):427–427.

9. Ehringer H, Hornykiewicz O. Distribution of noradrenaline and dopamine (3-hydroxytyramine) in the human brain and their behavior in diseases of the extrapyramidal system. Parkinsonism Relat Disord 1998;4(2):53–53.

10. Marras C, McDermott MP, Rochon PA, et al. Survival in Parkinson disease: thirteen-year follow-up of the DATATOP cohort. Neurology 2005;64(1):87–87.

11. Sweet RD, McDowell FH. Five years’ treatment of Parkinson’s disease with levodopa. Therapeutic results and survival of 100 patients. Annals of internal medicine 1975;83(4):456–456.

12. Barbeau A. High-level levodopa therapy in Parkinson’s disease: five years later. Trans Am Neurol Assoc 1974;99:160–163.

13. Barbeau A. Six years of high-level levodopa therapy in severely akinetic parkinsonian patients. Arch Neurol 1976;33(5):333–333.

14. De Pablo-Fernandez E, Tur C, Revesz T, Lees AJ, Holton JL, Warner TT. Association of Autonomic Dysfunction With Disease Progression and Survival in Parkinson Disease. JAMA Neurol 2017;74(8):970–970.

15. Hughes AJ, Daniel SE, Blankson S, Lees AJ. A clinicopathologic study of 100 cases of Parkinson’s disease. Arch Neurol 1993;50(2):140–140.

16. Sieber BA, Landis S, Koroshetz W, et al. Prioritized research recommendations from the National Institute of Neurological Disorders and Stroke Parkinson’s Disease 2014 conference. Ann Neurol 2014;76(4):469–469.

17. Kondo T, Mizuno Y, Japanese Istradefylline Study G. A long-term study of istradefylline safety and efficacy in patients with Parkinson disease. Clin Neuropharmacol 2015;38(2):41–41.

18. Giladi N, Gurevich T, Djaldetti R, et al. ND0612 (levodopa/carbidopa for subcutaneous infusion) in patients with Parkinson’s disease and motor response fluctuations: A randomized, placebo-controlled phase 2 study. Parkinsonism Relat Disord 2021;91:139–145.

19. Hauser RA, Espay, A.J., Lewitt, P., Ellenbogen, A., Isaacson, S. Pahwa, R., Stocchi, F., Visser, H., D’Souza, R. A phase III trial of IPX203 vs CD-LD in Parkinson’s disease patients with motor fluctuations (RISE-PD) (S16.010). Neurology 2022;98(May).

20. Soileau MJ, Aldred J, Budur K, et al. Safety and efficacy of continuous subcutaneous foslevodopa-foscarbidopa in patients with advanced Parkinson’s disease: a randomised, double-blind, active-controlled, phase 3 trial. Lancet Neurol 2022;21(12):1099–1099.

21. Krishna V, Fishman PS, Eisenberg HM, et al. Trial of Globus Pallidus Focused Ultrasound Ablation in Parkinson’s Disease. N Engl J Med 2023;388(8):683–683.

22. Papapetropoulos S, Liu W, Duvvuri S, Thayer K, Gray DL. Evaluation of D1/D5 Partial Agonist PF-06412562 in Parkinson’s Disease following Oral Administration. Neurodegener Dis 2018;18(5-6):262–269.

23. Riesenberg R, Werth J, Zhang Y, Duvvuri S, Gray D. PF-06649751 efficacy and safety in early Parkinson’s disease: a randomized, placebo-controlled trial. Ther Adv Neurol Disord 2020;13:1756286420911296.

24. McFarthing K, Rafaloff G, Baptista MAS, Wyse RK, Stott SRW. Parkinson’s Disease Drug Therapies in the Clinical Trial Pipeline: 2021 Update. Journal of Parkinson’s disease 2021;11(3):891–891.

25. Bidesi NSR, Vang Andersen I, Windhorst AD, Shalgunov V, Herth MM. The role of neuroimaging in Parkinson’s disease. J Neurochem 2021;159(4):660–660.

26. Lehericy S, Bardinet E, Poupon C, Vidailhet M, Francois C. 7 Tesla magnetic resonance imaging: a closer look at substantia nigra anatomy in Parkinson’s disease. Mov Disord 2014;29(13):1574–1574.

27. Ofori E, Pasternak O, Planetta PJ, et al. Increased free water in the substantia nigra of Parkinson’s disease: a single-site and multi-site study. Neurobiol Aging 2015;36(2):1097–1097.

28. Ofori E, Pasternak O, Planetta PJ, et al. Longitudinal changes in free-water within the substantia nigra of Parkinson’s disease. Brain 2015;138(Pt 8):2322–2331.

29. Du G, Lewis MM, Sen S, et al. Imaging nigral pathology and clinical progression in Parkinson’s disease. Mov Disord 2012;27(13):1636–1636.

30. Du G, Lewis MM, Sica C, et al. Distinct progression pattern of susceptibility MRI in the substantia nigra of Parkinson’s patients. Mov Disord 2018;33(9):1423–1423.

31. Du G, Lewis MM, Styner M, et al. Combined R2* and diffusion tensor imaging changes in the substantia nigra in Parkinson’s disease. Mov Disord 2011;26(9):1627–1627.

32. Manne S, Kondru N, Jin H, et al. alpha-Synuclein real-time quaking-induced conversion in the submandibular glands of Parkinson’s disease patients. Mov Disord 2020;35(2):268–268.

33. Manne S, Kondru N, Jin H, et al. Blinded RT-QuIC Analysis of alpha-Synuclein Biomarker in Skin Tissue From Parkinson’s Disease Patients. Mov Disord 2020;35(12):2230–2230.

34. Wang Z, Becker K, Donadio V, et al. Skin alpha-Synuclein Aggregation Seeding Activity as a Novel Biomarker for Parkinson Disease. JAMA Neurol 2020.

35. Siderowf A, Concha-Marambio L, Lafontant DE, et al. Assessment of heterogeneity among participants in the Parkinson’s Progression Markers Initiative cohort using alpha-synuclein seed amplification: a cross-sectional study. Lancet Neurol 2023;22(5):407–407.

36. Iwaki H, Leonard HL, Makarious MB, et al. Accelerating Medicines Partnership: Parkinson’s Disease. Genetic Resource. Mov Disord 2021;36(8):1795–1795.

37. Schwab RSE, A.C. Projection technique for evaluating surgery in Parkinson’s disease. In: Gillingham FJ, Donaldson MC, eds. Third symposium on Parkinson’s disease. Edinburgh: Livingston, 1969:152–157.

38. Jenkinson C, Fitzpatrick R, Peto V, Greenhall R, Hyman N. The Parkinson’s Disease Questionnaire (PDQ-39): development and validation of a Parkinson’s disease summary index score. Age Ageing 1997;26(5):353–353.

39. Goetz CG, Fahn S, Martinez-Martin P, et al. Movement Disorder Society-sponsored revision of the Unified Parkinson’s Disease Rating Scale (MDS-UPDRS): Process, format, and clinimetric testing plan. Mov Disord 2007;22(1):41–41.

40. Doty RL, Shaman P, Dann M. Development of the University of Pennsylvania Smell Identification Test: a standardized microencapsulated test of olfactory function. Physiol Behav 1984;32(3):489–489.

41. Nasreddine ZS, Phillips NA, Bedirian V, et al. The Montreal Cognitive Assessment, MoCA: a brief screening tool for mild cognitive impairment. J Am Geriatr Soc 2005;53(4):695–695.

42. Johns MW. A new method for measuring daytime sleepiness: the Epworth sleepiness scale. Sleep 1991;14(6):540–540.

43. Du G, Wang E, Sica C, et al. Dynamics of Nigral Iron Accumulation in Parkinson’s Disease: From Diagnosis to Late Stage. Mov Disord 2022;37(8):1654–1654.

44. Lewis MM, Harkins E, Lee EY, et al. Clinical Progression of Parkinson’s Disease: Insights from the NINDS Common Data Elements. Journal of Parkinson’s disease 2020;10(3):1075–1075.

45. Sterling NW, Wang M, Zhang L, et al. Stage-dependent loss of cortical gyrification as Parkinson disease “unfolds”. Neurology 2016;86(12):1143–1143.

46. Doty RL. The smell dientification test administration manual. New Jersey: Sensonics, Inc; 1995.

47. Cerri S, Mus L, Blandini F. Parkinson’s Disease in Women and Men: What’s the Difference? J Parkinsons Dis 2019;9(3):501–501.

48. Haaxma CA, Bloem BR, Borm GF, et al. Gender differences in Parkinson’s disease. J Neurol Neurosurg Psychiatry 2007;78(8):819–819.

49. Sieurin J, Andel R, Tillander A, Valdes EG, Pedersen NL, Wirdefeldt K. Occupational stress and risk for Parkinson’s disease: A nationwide cohort study. Mov Disord 2018;33(9):1456–1456.

50. Rafferty MR, Schmidt PN, Luo ST, et al. Regular Exercise, Quality of Life, and Mobility in Parkinson’s Disease: A Longitudinal Analysis of National Parkinson Foundation Quality Improvement Initiative Data. Journal of Parkinson’s disease 2017;7(1):193–193.

51. Baba Y, Putzke JD, Whaley NR, Wszolek ZK, Uitti RJ. Gender and the Parkinson’s disease phenotype. J Neurol 2005;252(10):1201–1201.

52. Colombo D, Abbruzzese G, Antonini A, et al. The “gender factor” in wearing-off among patients with Parkinson’s disease: a post hoc analysis of DEEP study. ScientificWorldJournal 2015;2015:787451.

53. Latourelle JC, Beste MT, Hadzi TC, et al. Large-scale identification of clinical and genetic predictors of motor progression in patients with newly diagnosed Parkinson’s disease: a longitudinal cohort study and validation. Lancet Neurol 2017;16(11):908–908.

54. Sperens M, Georgiev D, Eriksson Domellof M, Forsgren L, Hamberg K, Hariz GM. Activities of daily living in Parkinson’s disease: Time/gender perspective. Acta Neurol Scand 2020;141(2):168–168.

55. Khedr EM, El Fetoh NA, Khalifa H, Ahmed MA, El Beh KM. Prevalence of non motor features in a cohort of Parkinson’s disease patients. Clin Neurol Neurosurg 2013;115(6):673–673.

56. Locascio JJ, Corkin S, Growdon JH. Relation between clinical characteristics of Parkinson’s disease and cognitive decline. J Clin Exp Neuropsychol 2003;25(1):94–94.

57. Cholerton B, Johnson CO, Fish B, et al. Sex differences in progression to mild cognitive impairment and dementia in Parkinson’s disease. Parkinsonism Relat Disord 2018;50:29–36.

58. Wickremaratchi MM, Ben-Shlomo Y, Morris HR. The effect of onset age on the clinical features of Parkinson’s disease. Eur J Neurol 2009;16(4):450–450.

59. Pagano G, Ferrara N, Brooks DJ, Pavese N. Age at onset and Parkinson disease phenotype. Neurology 2016;86(15):1400–1400.

60. Raket LL, Oudin Astrom D, Norlin JM, Kellerborg K, Martinez-Martin P, Odin P. Impact of age at onset on symptom profiles, treatment characteristics and health-related quality of life in Parkinson’s disease. Sci Rep 2022;12(1):526.

61. Bega D, Kim S, Zhang Y, et al. Predictors of Functional Decline in Early Parkinson’s Disease: NET-PD LS1 Cohort. J Parkinsons Dis 2015;5(4):773–773.

62. Marras C, Rochon P, Lang AE. Predicting motor decline and disability in Parkinson disease: a systematic review. Arch Neurol 2002;59(11):1724–1724.

63. Perneczky R, Kempermann G, Korczyn AD, et al. Translational research on reserve against neurodegenerative disease: consensus report of the International Conference on Cognitive Reserve in the Dementias and the Alzheimer’s Association Reserve, Resilience and Protective Factors Professional Interest Area working groups. BMC Med 2019;17(1):47.

64. Kostic V, Przedborski S, Flaster E, Sternic N. Early development of levodopa-induced dyskinesias and response fluctuations in young-onset Parkinson’s disease. Neurology 1991;41(2 (Pt 1)):202–205.

65. Ku S, Glass GA. Age of Parkinson’s disease onset as a predictor for the development of dyskinesia. Mov Disord 2010;25(9):1177–1177.

66. Huang X, Lewis MM, Van Scoy LJ, et al. The D1/D5 Dopamine Partial Agonist PF-06412562 in Advanced-Stage Parkinson’s Disease: A Feasibility Study. Journal of Parkinson’s disease 2020;10(4):1515–1515.

67. Lewis MM, Van Scoy LJ, De Jesus S, et al. Dopamine D(1) Agonists: First Potential Treatment for Late-Stage Parkinson’s Disease. Biomolecules 2023;13(5).

68. Mailman RB, Yang Y, Huang X. D1, not D2, dopamine receptor activation dramatically improves MPTP-induced parkinsonism unresponsive to levodopa. European journal of pharmacology 2021;892:173760.

